# Increased Interregional Travel to Shopping Malls and Restaurants in Response to Differential COVID-19 Restrictions in the Greater Toronto Area

**DOI:** 10.1101/2021.04.23.21255959

**Authors:** Jean-Paul R. Soucy, Amir Ghasemi, Shelby L. Sturrock, Isha Berry, Sarah A. Buchan, Derek R. MacFadden, Nick Daneman, Nicholas Gibb, Kevin A. Brown

## Abstract

**Background:** In the fall of 2020, the government of Ontario, Canada adopted a 5-tier, regional framework of public health measures for the COVID-19 pandemic. During the second wave of COVID-19 in Ontario, the urban core of the Greater Toronto Area (Toronto and Peel) were the first regions in the province to enter the highest restriction tier (“lockdown”) on November 23, 2020, which closed restaurants to in-person dining and limited non-essential businesses, including shopping malls, to curbside pickup. The peripheral regions of the Greater Toronto Area (York, Durham, Halton) would not enter lockdown until later the following month. In this analysis, we examine whether the implementation of differentially timed restrictions in a highly interconnected metropolitan area led to increased interregional travel, potentially driving further transmission of SARS-CoV-2.

**Methods:** We used anonymized smartphone data to estimate the number of visits by residents of regions in the urban core to shopping malls and restaurants in peripheral regions in the week before compared to the week after the November 23 lockdown.

**Results:** Residents of Toronto and Peel took fewer trips to shopping malls and restaurants in the week following lockdown. This was entirely driven by reductions in visits within the locked down regions themselves, as there was a significant increase in trips to shopping malls in peripheral regions by these residents in the same period (Toronto: +40.7%, Peel: +65.5%). Visits to restaurants in peripheral regions also increased slightly (Toronto: +6.3%, Peel: +11.8%).

**Discussion:** Heterogeneous restrictions may undermine lockdowns in the urban core as well as driving residents from zones of higher transmission to zones of lower transmission. These concerns are likely generalizable to other major metropolitan areas, which often comprise interconnected but administratively independent regions.

## Introduction

In the fall of 2020, the government of Ontario, Canada adopted a 5-tier, regional framework of public health measures for the COVID-19 pandemic in its 34 public health regions. ^1^ The goal of non-pharmaceutical interventions is to suppress transmission by reducing contact rates, which can be indirectly assessed using mobility data. Five of the 6 most populous health regions are located in the Greater Toronto Area: Toronto (3.0 million), Peel (1.5 million), York (1.2 million), Durham (0.7 million) and Halton (0.6 million) (Supplementary Figure 1). The urban core of Toronto and Peel is a perpetual hotspot for COVID-19^2^ and remains highly interconnected with the peripheral regions of York, Durham and Halton.

During the second wave of COVID-19 in Ontario, Toronto and Peel were the first regions in the province to enter the highest restriction tier (“lockdown”) on November 23, 2020, which closed restaurants to in-person dining and limited non-essential businesses, including shopping malls, to curbside pickup. The peripheral region of York entered lockdown on December 14, followed by the rest of the province (including Durham and Halton) on December 26. In this analysis, we examine whether the implementation of differentially timed restrictions in a highly interconnected metropolitan area led to increased interregional travel, potentially driving further transmission of SARS-CoV-2.

## Methods

We used anonymized smartphone data from Veraset (www.veraset.com) representing approximately 1% of the Canadian population to analyze patterns of travel by residents of regions in the urban core (Toronto and Peel) to shopping malls and restaurants in peripheral regions in the week before compared to the week after the November 23 lockdown. Restaurants and shopping malls are both important settings for transmission risk. ^3,4^ A device’s home region for a given calendar month was identified as the region where the device spent most of its time over the course of that month. The proportion of devices in the dataset that visited malls or restaurants was multiplied by the population of the region (2019 estimates) ^5^ to estimate the actual number of visitors. We also measured visits by residents of Toronto and Peel to shopping malls in York relative to a baseline calculated for each day of the week from January 1 to February 5, 2020.

Ethics approval was obtained from the University of Toronto Research Ethics Board through the Ontario COVID-19 Modelling Consensus Table.

## Results

Residents of Toronto and Peel took fewer trips to shopping malls and restaurants in the week following lockdown (Table 1). This was entirely driven by reductions in visits within the locked down regions themselves, as there was a significant increase in trips to shopping malls in eripheral regions by these residents in the same period (Toronto: +40.7%, Peel: +65.5%). However, visits to peripheral regions were still well below historical averages (Figure 1). Visits to restaurants in peripheral regions also increased slightly (Toronto: +6.3%, Peel: +11.8%).

**Table 1.**
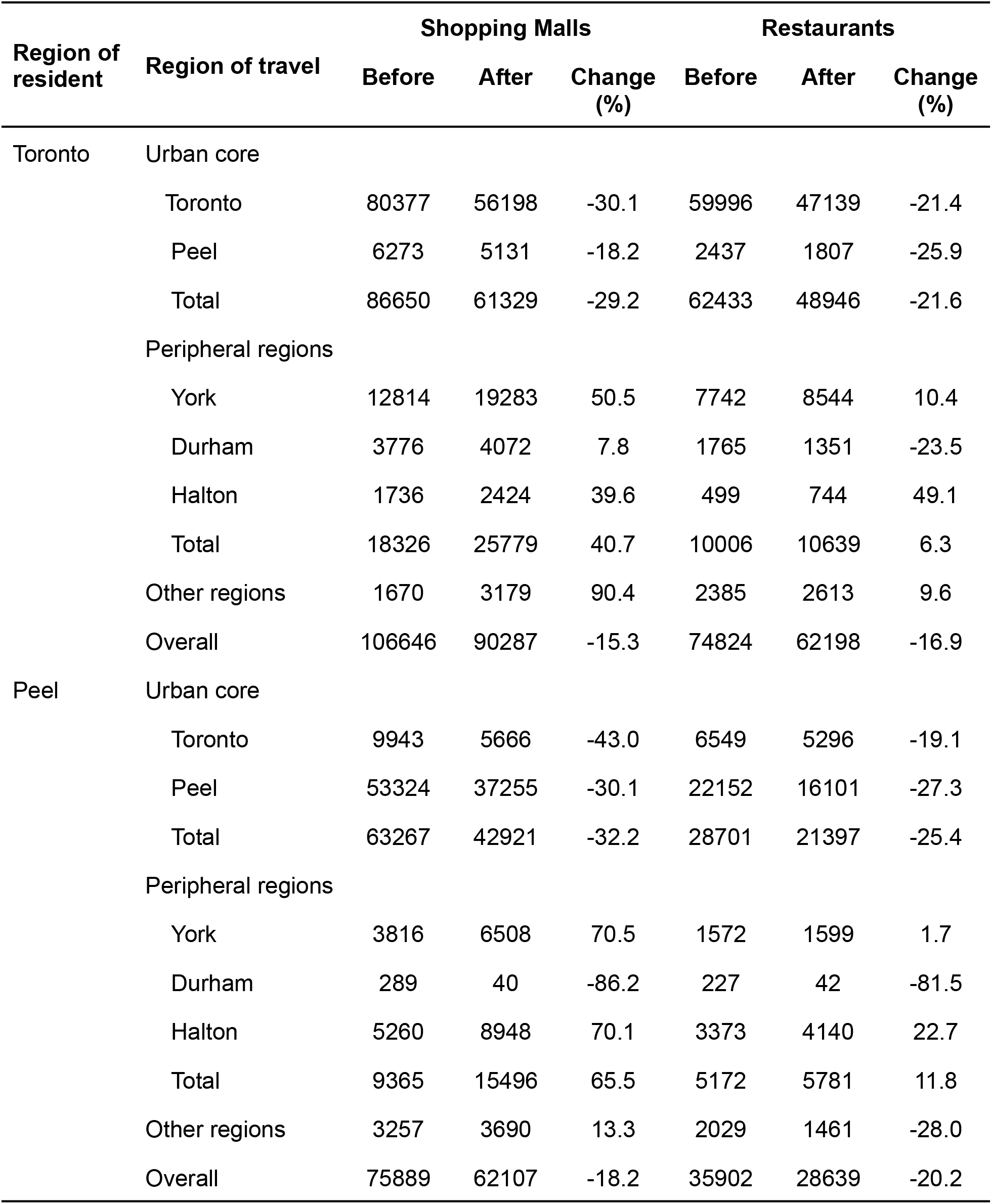
Estimated number of Toronto and Peel residents visiting shopping malls and restaurants in other regions of Ontario, one week before and after lockdown (November 23, 2020).

**Figure 1.**
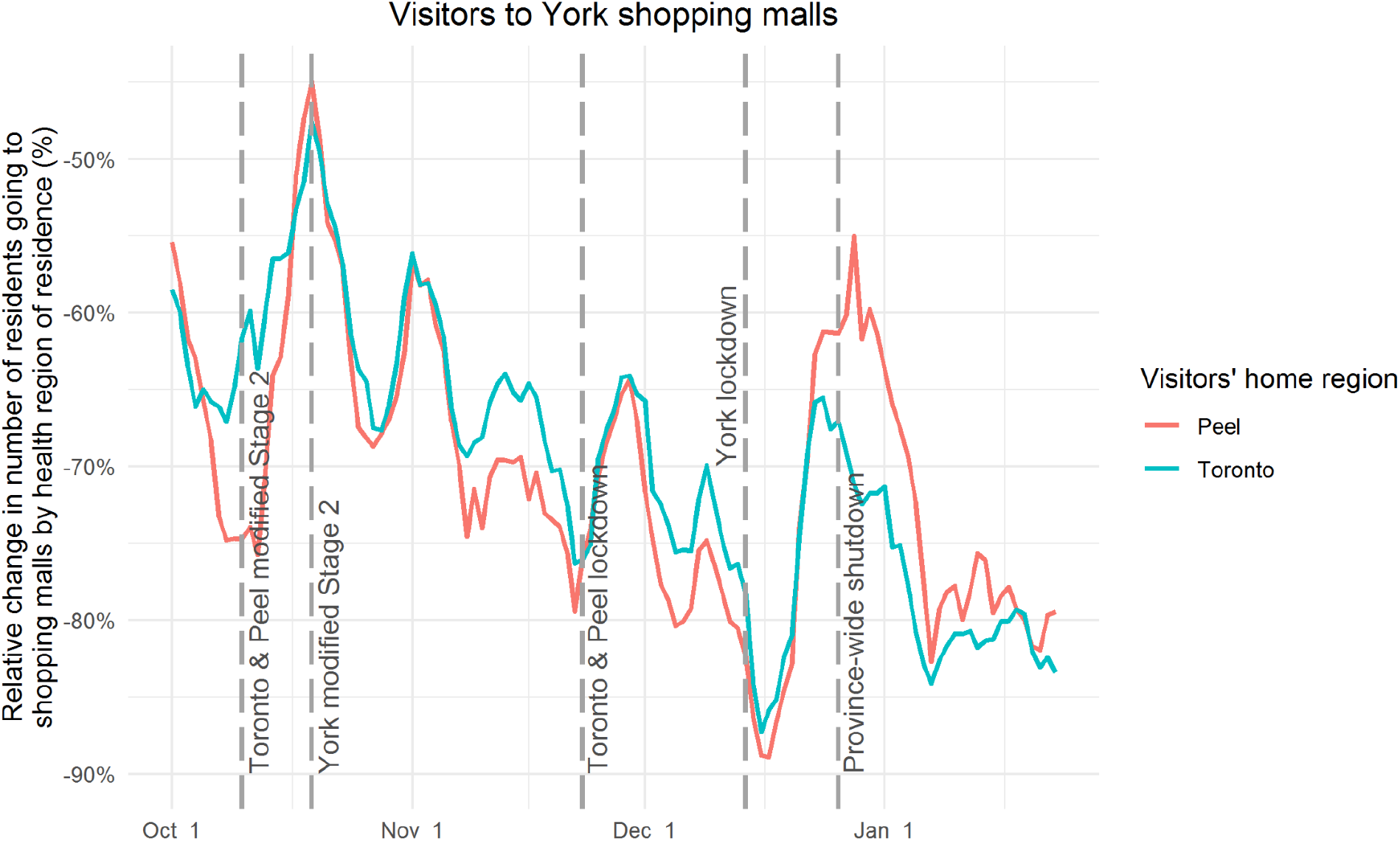
Visits from Toronto and Peel residents to shopping malls in York, relative to a baseline calculated for each day of the week from January 1 to February 5, 2020. Toronto and Peel entered lockdown on November 23, 2020. York entered lockdown on December 14, 2020. Other regions entered lockdown on December 26, 2020.

## Discussion

Lockdowns in the urban core reduced overall visits to shopping malls and restaurants but increased visits to peripheral regions, where restrictions permitted indoor dining and shopping for non-essential businesses. These heterogeneous restrictions may lead to unintended consequences, undermining lockdowns in the urban core and driving residents from zones of higher transmission to zones of lower transmission. In this analysis, shopping malls appear particularly likely to be affected by regional variation in restrictions. Regional non-pharmaceutical intervention frameworks could avoid this by implementing restrictions spanning both the core and periphery of urban areas or by using interregional travel restrictions. These concerns are likely generalizable to other major metropolitan areas, which often comprise interconnected but administratively independent regions. ^6^

## Data Availability

The dataset used to support the conclusions of this paper is presented in Table 1. The time series presented in Figure 1 are available upon request.

## Supplementary material

**Supplementary figure 1.**
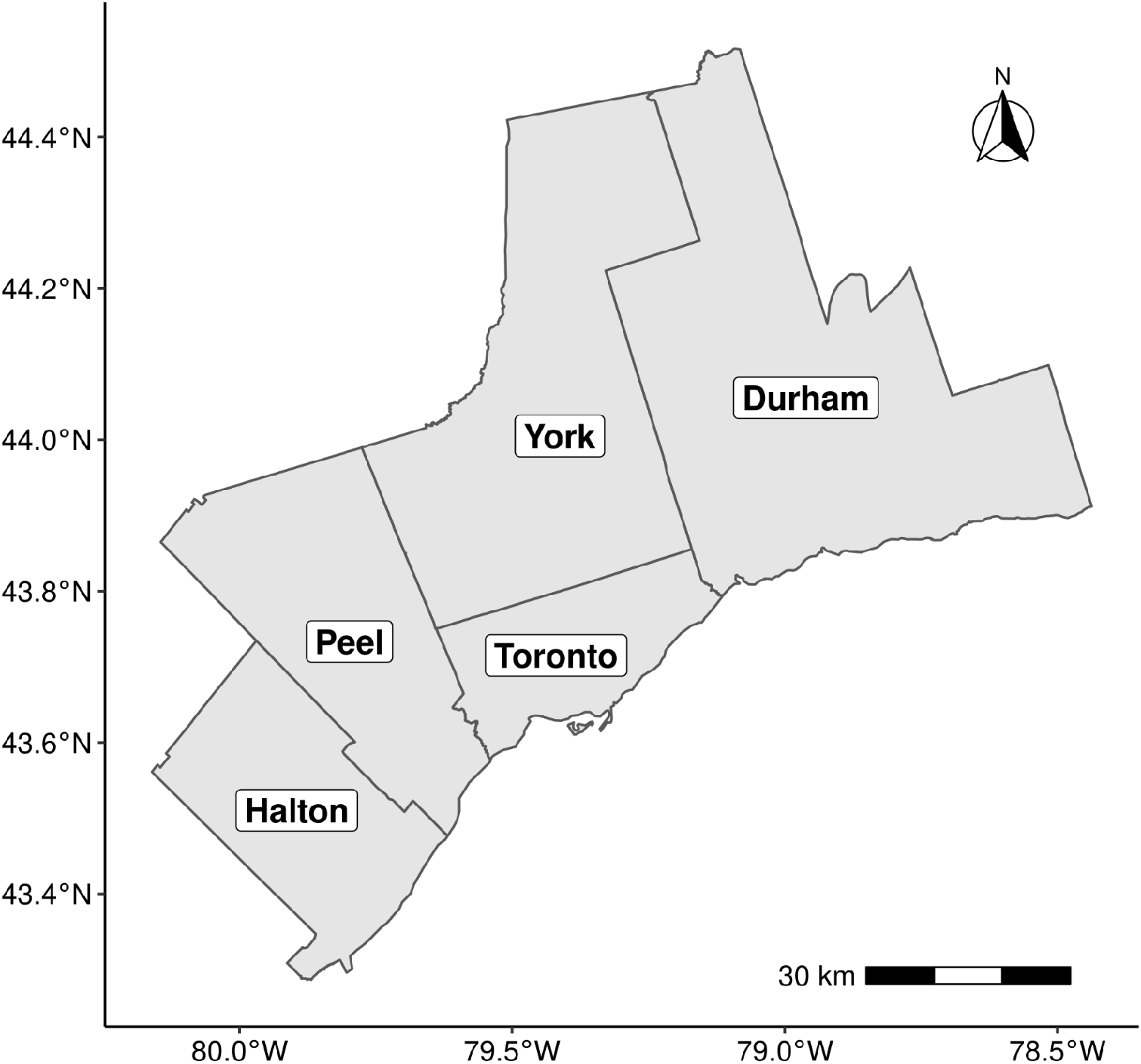
Map of the five regions comprising the Greater Toronto Area.

## Notes

### Competing Interest Statement

The authors have declared no competing interest.

### Funding Statement

This study received no external funding.

### Author Declarations

Ethics approval was obtained from the University of Toronto Research Ethics Board through the Ontario COVID-19 Modelling Consensus Table, a provincial working group developing evidence for the COVID-19 response that operates with the support of the Ontario Ministry of Health, Ontario Health and Public Health Ontario.

